# Integrated Protein Network Analysis of Whole Exome Sequencing of Severe Preeclampsia

**DOI:** 10.1101/2020.11.23.20236885

**Authors:** Jessica Schuster, George A. Tollefson, Valeria Zarate, Anthony Agudelo, Joan Stabila, Ashok Ragavendran, James Padbury, Alper Uzun

## Abstract

Preeclampsia is a hypertensive disorder of pregnancy, which complicates up to 15 % of US deliveries. It is an idiopathic disorder with complex disease genetics associated with several different phenotypes. We sought to determine if the genetic architecture of preeclampsia can be described by clusters of patients with variants in genes in shared protein interaction networks. We performed a case-control study using whole exome sequencing on early onset preeclamptic mothers with severe features and control mothers with uncomplicated pregnancies. The study was conducted at Women & Infants Hospital of Rhode Island (WIH). A total of 143 patients were enrolled, 61 women with early onset preeclampsia with severe features based on ACOG criteria, and 82 control women at term, matched for race and ethnicity. The main outcomes are variants associated with severe preeclampsia and demonstration of the genetic architecture of preeclampsia. A network analysis and visualization tool, Proteinarium, was used to confirm there are clusters of patients with shared gene networks associated with severe preeclampsia. The majority of the sequenced patients appear in two significant clusters. We identified one case dominant and one control dominant cluster. Thirteen genes were unique to the case dominated cluster. Among these genes, LAMB2, PTK2, RAC1, QSOX1, FN1, and VCAM1 have known associations with the pathogenic mechanisms of preeclampsia. Using the exome-wide sequence variants, combined with these 13 identified network genes, we generated a polygenetic risk score for severe preeclampsia with an AUC of 0.57. Using bioinformatic analysis, we were able to identify subsets of patients with shared protein interaction networks, thus confirming our hypothesis about the genetic architecture of preeclampsia. The unique genes identified in the cluster associated with severe preeclampsia were able to increase the predictive power of the polygenic risk score.

## Introduction

Preeclampsia is a hypertensive disorder of pregnancy. It is associated with a higher risk of hypertension and cardiovascular disease later in life. Women who had preeclampsia have a twofold increased risk of death from cardiovascular diseases ^1, 2^. There is evidence that preeclampsia originates in part from genetic causes that include contributions from the maternal, paternal and fetal genome ^3-7^. The role of genetics in preeclampsia is supported by family-based observations ^8, 9^ with more than 100 studies showing a 2- to 5-fold increased risk among family members of affected women ^10-15^. The heritability of preeclampsia is up to 52% ^8, 16^. The recurrence risk for preeclampsia in the daughters of either eclamptic or preeclamptic patients is 20-40% ^17, 18^. However, there is no current consensus among the published results in regards to associated genes and the pathogenesis of disease.

Genetic risk for most complex diseases involves the interaction of multiple genes in discrete networks and pathways ^19^. Although complex diseases show increased recurrence risk in families, they do not follow a simple Mendelian pattern of inheritance ^20^. Computational methods have been used to analyze the network of genes that are linked to a variety of disorders like autism and to find biological subnetworks due to the genetic heterogeneity of the disease ^21^. There are several studies employing computational methods to identify important genes associated with hypertension. Ran et al analyzed protein-protein interaction (PPI) network topology and molecular connectivity between protein pathways to identify associations with hypertension ^22^. Researchers developed a machine-learning algorithm to predict novel hypertension associated genes^23^.

We hypothesize that the genetic architecture of complex diseases like preeclampsia is described by clusters of patients with variants in genes in shared protein interaction networks. We sought to test this hypothesis using whole exome sequencing in carefully selected patients with severe preeclampsia. We compared variants identified in women with early onset, idiopathic preeclampsia with term controls without personal or family history of pregnancy related hypertensive disorders. We built and implemented *Proteinarium*, a multi-sample, PPI tool, to identify clusters of patients with shared PPI networks.

## Methods

### Study population

Women & Infants Hospital of Rhode Island (WIH) is the only provider of high-risk perinatal services in Rhode Island, northeastern Connecticut and southeastern Massachusetts. We used this population-based service to enroll preeclamptic mothers with early onset, severe features, based on ACOG criteria, as well as term mothers with no history of preeclampsia ^24^.

This case/control study was approved by the Institutional Review Board of WIH (Project ID: WIH 16-0031). Between the years 2016-2020, we reviewed the records of all early-onset preeclamptic mothers with severe features delivering < 34 weeks. Following informed consent, we asked explicit questions about preeclampsia in mother, grandmother, first order relatives and also paternal relatives. Clinical history, with an emphasis on additional risk factors including medical illnesses and drug use was recorded. Hypertensive disorders include a broad range of different phenotypes. Again, in order to leverage the likelihood of genetic discovery associated with preeclampsia, we excluded mothers with personal or family history of other hypertensive disorders. Controls were mothers who delivered ≥ 37 weeks’ gestation for whom the formal genetic interview revealed no history of preterm birth or pregnancy related hypertensive disorders on either the maternal or paternal side of the pedigree. A total of 143 patients were enrolled, 61 women with early onset preeclampsia with severe features, and 82 control women at term, matched for race and ethnicity.

### Whole Exome Sequencing

Residual maternal whole blood was obtained from each mother and stored at -80°C. Samples were sent to an outside facility for whole exome sequencing that was blind to disease status. The library was sequenced on an Illumina HiSeq 4000 using 150 bp paired-end protocols.

### Sequence Data

For variant discovery we used the Gene Analysis Tool Kit (GATK) V4 to analyze the sequence reads ^25^. Haplotype caller was applied for variant detection ^26^. Variants were flagged as low quality and filtered using established metrics: if three or more variants were detected within 10bp; if four or more alignments mapped to different locations equally well; if coverage was less than ten reads; if quality score < 30; if low quality for a particular sequence depth (variant confidence/unfiltered depth < 1.5); and if strand bias was observed (Phred-scaled p-values using Fisher’s Exact Test > 200).

### Genotype Testing

To identify variants that were differentially abundant between cases and controls, we used a Markov Chain Monte Carlo (MCMC) Fisher Exact Test to compare the frequency of the homozygous reference, homozygous alternative, and the heterozygous genotypes between cases and controls. Eigenstrat detected no significant population stratification ^27^.

### Variant Annotation

We applied a strict filter-based annotation using ANNOVAR ^28^. We identified deleterious variants with Polyphen 2 HDIV, SIFT and CADD ^29-32^. We used the following thresholds: Polyphen 2 HDIV prediction if a change is damaging (>=0.957), a SIFT score (<0.05), a CADD score >15, and minor allele frequency (MAF) <0.05 from the 1000 Genome Project^32^.

### Network Analysis

We hypothesized that the genetic architecture underlying complex disorders is best explained by subsets of patients with variants in shared networks and pathways sufficient to express the phenotype. To analyze our whole exome sequencing data, we implemented *Proteinarium*, our multi-sample PPI analysis and visualization tool ^1^. We determined the genetic similarity between the clusters identified using separation testing. Combining genome wide variants with the unique genes identified by this network analysis, we generated a polygenic risk score prediction model. These analyses are explained in detail in the Supplementary Methods.

## Results

The clinical characteristics and the race/ethnicity distribution of the patients are shown in Table 1. As can be seen from Table 1, gestational age at delivery, systolic blood pressure, frequency of proteinuria, impaired liver function, thrombocytopenia, cerebral visual symptoms and fetal growth retardation were all significantly different between the groups, which was expected by our definition of severe preeclampsia.

**Table 1.** Clinical characteristics of patients. Mean + SD.

| **Categories** | **case (n=61)** | **control (n=82)** |
| --- | --- | --- |
| **Gestational age of delivery and life style** |  |  |
| *Age (mean)* | 29.1 ± 5.0 | 29.4 ± 5.3 |
| *Grava (mean)* | 2.1 ± 1.2 | 2.5 ± 1.6 |
| *Job strenuous (%)* | 26.2% | 28.0% |
| *Obesity (%)* | 31.1% | 23.1% |
| **Race/Ethnicity** |  |  |
| *African_American (%)* | 9.8% | 4.8% |
| *Asian (%)* | 3.2% | 3.6% |
| *Caucasian (%)* | 55.7% | 56.1% |
| *Hispanic (%)* | 22.9% | 28.0% |
| *Native_American (%)* | 1.6% | 1.2% |
| *Other_Racial_ID (%)* | 6.5% | 6.1% |
| **Abnormal laboratory values** |  |  |
| *Systolic_bp (mean, mmHg)* | 170.8 ± 14.4 | 117.6 ± 9.6 |
| *Proteinuria (%)* | 65.5% | 0.00% |
| *Impaired_liver_function (%)* | 55.7% | 2.4% |
| *Thrombocytopenia (%)* | 14.7% | 0.0% |
| *Cerebral_visual_symptoms (%)* | 55.7% | 0.0% |
| *Fetal Growth Restriction (FGR) (%)* | 29.5% | 2.4% |
| **Preterm delivery** |  |  |
| *Preterm_delivery_before_34_weeks_for_sPEC (%)* | 55.7% | 0.0% |
| *Preterm_delivery_before_37_weeks (%)* | 60.6% | 3.6% |

High quality sequence data with a Phred score ≥30 from well-balanced pools with over 19,000,000 reads/patient, 40X average depth of coverage, with more than 80% of sequence reads with at least 20X coverage were observed. We identified a total of 528,630 variants including 187,915 exonic variants. The work flow for the univariate analysis is shown in Figure 1. After application of the initial filters for coverage and variant pathogenicity, there were 8,867 predicted deleterious variants (available at <u>Online_Supplemental_Table 1</u>). Among these, 21 variants were nominally associated with preeclampsia by genotype testing. All were non-synonymous, exonic variants (Table 2). Nonetheless, none of these variants met genome-wide significance after correction for multiple comparison testing.

**Figure 1.**
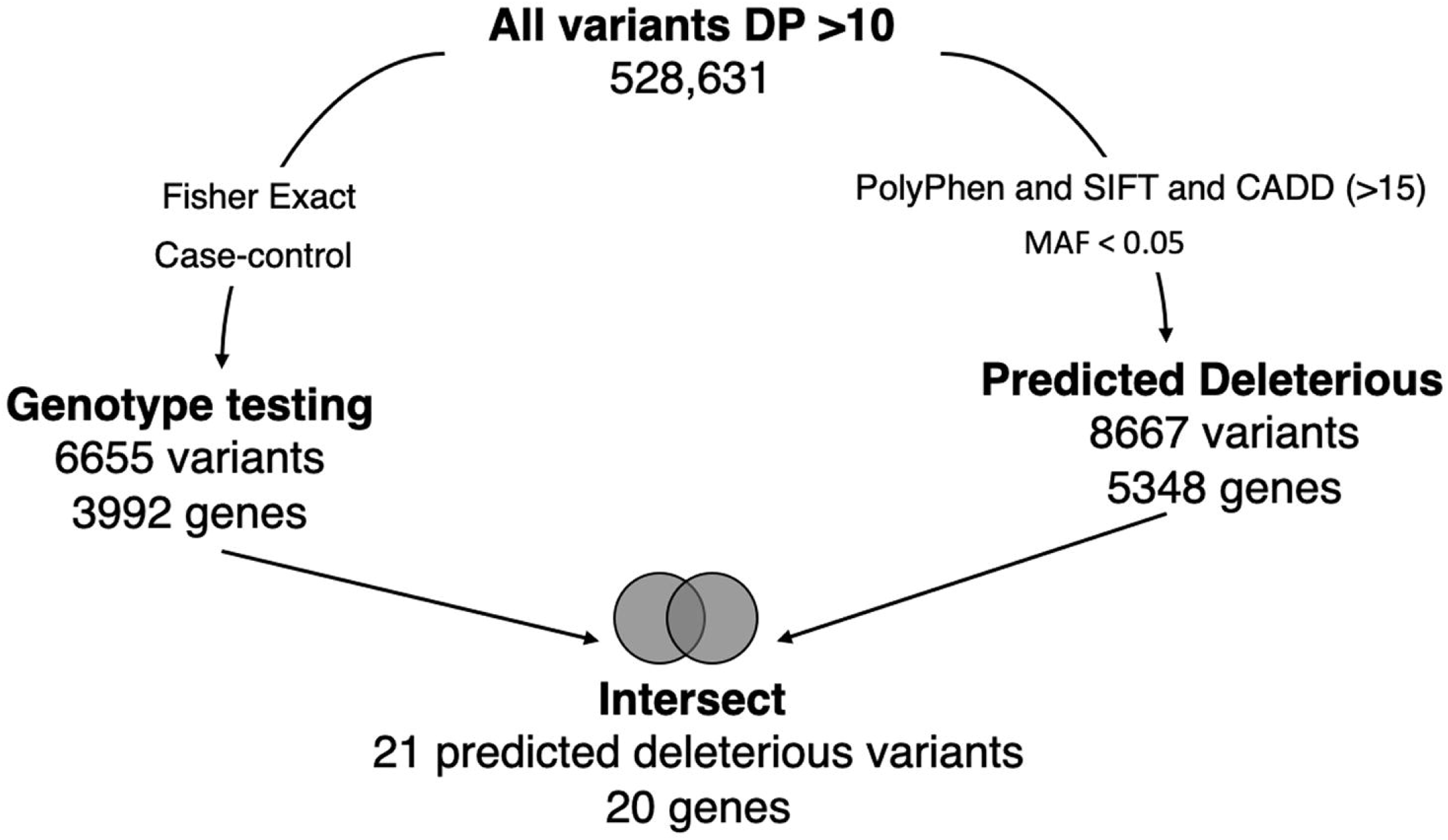
Figure shows the univariate work flow for analysis of the whole exome sequencing results.

**Table 2.** Pathogenic, nominally significant (based on genotype testing, p <0.05) gene variants identified by univariate analysis. Genomic positions are based on Human Feb. 2009 (GRCh37/hg19) Assembly.

| Chr | Pos | Gene | HGNC ID | SNP | p value | Polyphen 2_HDIV | SIFT | CADD |
| --- | --- | --- | --- | --- | --- | --- | --- | --- |
| 1 | 97770920 | DPYD | 3012 | rs1801160 | 0.032 | 0.998 | 0 | 23.5 |
| 1 | 104117921 | AMY2B | 478 | rs140978983 | 0.035 | 1 | 0 | 26.1 |
| 1 | 109446750 | GPSM2 | 29501 | rs61754640 | 0.022 | 0.994 | 0.02 | 19.3 |
| 1 | 226125385 | LEFTY2 | 3122 | rs2295418 | 0.022 | 1 | 0 | 16.6 |
| 2 | 69177269 | GKN2 | 24588 | rs62133344 | 0.036 | 1 | 0 | 18.5 |
| 2 | 70504399 | PCYOX1 | 20588 | rs34041544 | 0.030 | 1 | 0.01 | 26.4 |
| 2 | 179486345 | TTN | 12403 | rs114331773 | 0.017 | 1 | 0 | 15.7 |
| 2 | 179666982 | TTN | 12403 | rs35683768 | 0.022 | 0.999 | 0 | 15.7 |
| 6 | 76024704 | FILIP1 | 21015 | rs62415695 | 0.009 | 1 | 0.01 | 15.4 |
| 6 | 84904604 | CEP162 | 21107 | rs17790493 | 0.024 | 1 | 0 | 15.9 |
| 7 | 103130222 | RELN | 9957 | rs73714410 | 0.034 | 0.972 | 0.02 | 27.9 |
| 12 | 124221796 | ATP6V0A2 | 18481 | rs74922060 | 0.010 | 1 | 0.03 | 23.0 |
| 13 | 113750905 | MCF2L | 14576 | rs140657264 | 0.024 | 0.999 | 0 | 26.6 |
| 16 | 29825022 | PRRT2 | 30500 | rs76335820 | 0.043 | 0.995 | 0.02 | 18.4 |
| 17 | 34311387 | CCL14 | 10612 | rs16971802 | 0.047 | 0.974 | 0.02 | 16.2 |
| 17 | 37321347 | ARL5C | 31111 | rs9912267 | 0.028 | 1 | 0 | 18.6 |
| 18 | 28604374 | DSC3 | 3037 | rs35630063 | 0.021 | 1 | 0 | 21.1 |
| 19 | 56249615 | NLRP9 | 22941 | rs80009430 | 0.012 | 1 | 0 | 16.0 |
| 20 | 3641868 | GFRA4 | 13821 | rs146579049 | 0.017 | 1 | 0 | 18.3 |
| 20 | 36954724 | BPI | 1095 | rs5743523 | 0.008 | 0.998 | 0.02 | 15.5 |
| 22 | 31494813 | SMTN | 11126 | rs80055673 | 0.011 | 1 | 0.03 | 18.7 |

*Proteinarium* was used to identify clusters of patients with shared networks associated with severe preeclampsia and the resulting dendrogram is shown in Figure 2A. Out of the 143 patients sequenced, 129 patients were assigned to two statistically significant clusters. (*p<* 0.0001). The inset in Figure 2A shows the number of cases and controls in each cluster. Cluster A had significantly more cases than controls, containing 47 of the 61 case subjects. The layered network for the case-dominated Cluster A is shown in Figure 2B. There are 13 genes which are unique to Cluster A highlighted in red in the layered network graph. Most have defined functional roles or implications for preeclampsia, Table 3. Cluster B had significantly more controls than cases, including 61 of the 82 subjects. The layered network for Cluster B is shown in Figure 2C. The unique genes from the layered network graph of Cluster B, shown in blue, are listed in Supplemental Table 2. When we compared the sequence data of the samples not assigned to clusters with those that were assigned, we did not find significant differences in the average depth of coverage. Likewise, there were no significant differences in clinical/phenotypic characteristics when comparing the subjects in the significant clusters with the subjects that were not in these clusters (data not shown).

**Figure 2.**
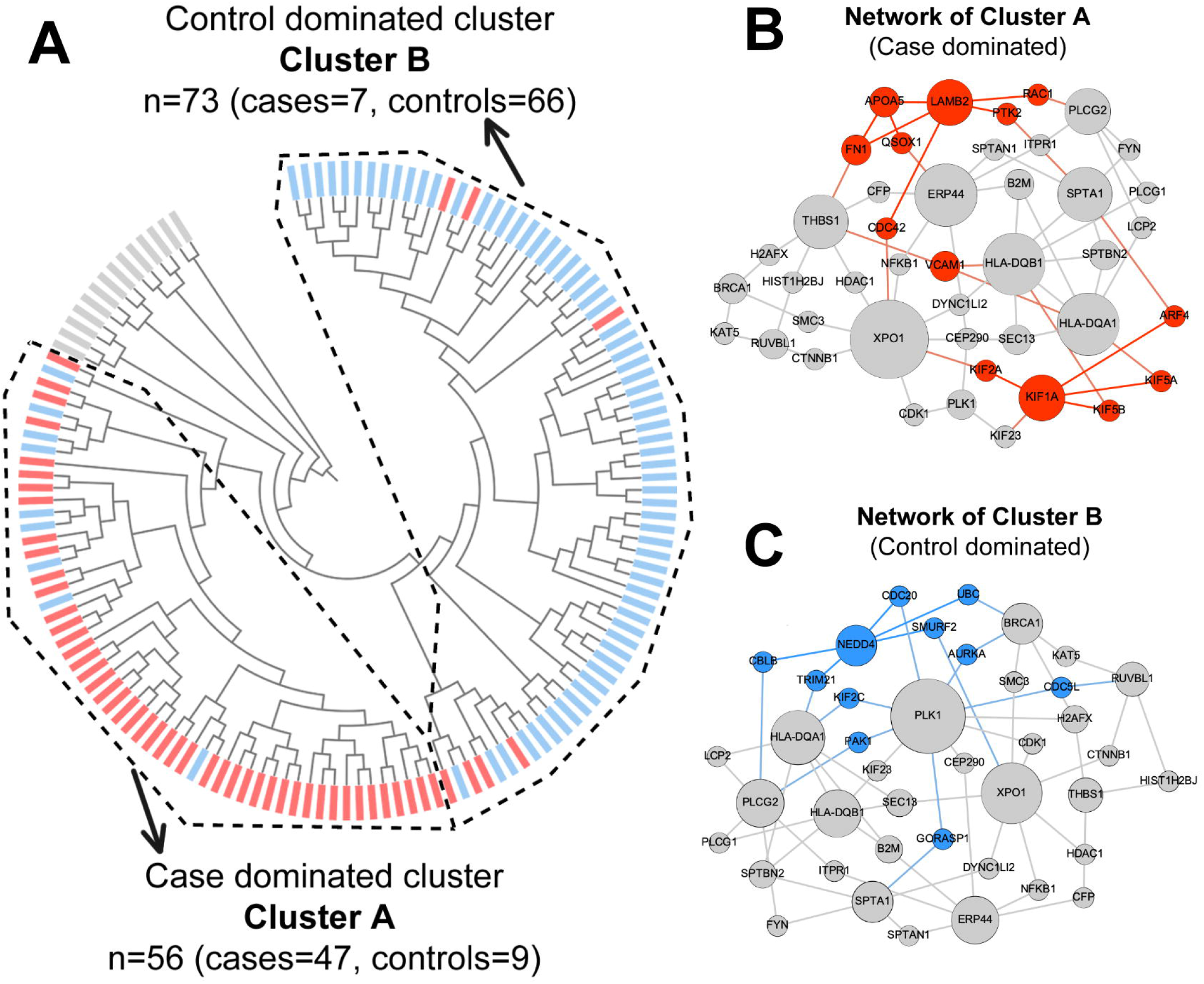
**A)** Dendrogram shows statistically significant (p <0.05) clusters of patients. Case dominated cluster (Cluster A) and control dominated cluster (Cluster B) are presented by dashed lines. Cases are represented in red and controls are represented in blue color. **B**) Layered network graphs for the case dominated cluster A are presented. Unique genes of cluster A are in red color. **C)** Layered network graphs for the control dominated cluster B are presented. Unique genes of cluster B are in blue color.

**Table 3.**
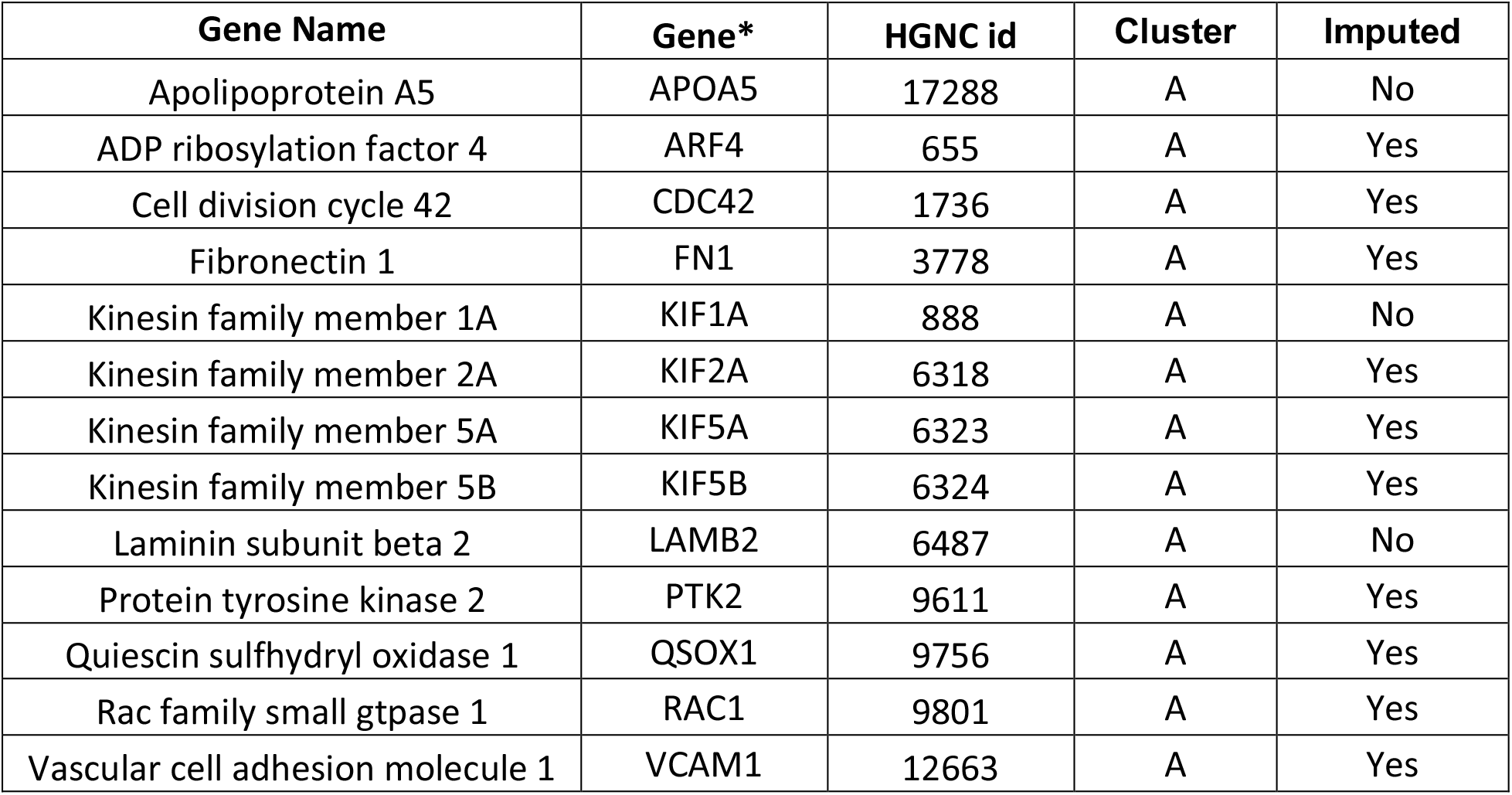
Unique genes from case dominated cluster (Cluster A). *Genes alphabetically ordered.

| Gene Name | Gene* | HGNC id | Cluster | Imputed |
| --- | --- | --- | --- | --- |
| Apolipoprotein A5 | APOA5 | 17288 | A | No |
| ADP ribosylation factor 4 | ARF4 | 655 | A | Yes |
| Cell division cycle 42 | CDC42 | 1736 | A | Yes |
| Fibronectin 1 | FN1 | 3778 | A | Yes |
| Kinesin family member 1A | KIF1A | 888 | A | No |
| Kinesin family member 2A | KIF2A | 6318 | A | Yes |
| Kinesin family member 5A | KIF5A | 6323 | A | Yes |
| Kinesin family member 5B | KIF5B | 6324 | A | Yes |
| Laminin subunit beta 2 | LAMB2 | 6487 | A | No |
| Protein tyrosine kinase 2 | PTK2 | 9611 | A | Yes |
| Quiescin sulfhydryl oxidase 1 | QSOX1 | 9756 | A | Yes |
| Rac family small gtpase 1 | RAC1 | 9801 | A | Yes |
| Vascular cell adhesion molecule 1 | VCAM1 | 12663 | A | Yes |

The comparison of the unique genes from the case and the control dominated clusters revealed a positive separation score, confirming that the layered PPI networks of these two patient subgroups exist in distinct areas of the interactome. We ran GO term analysis using DAVID software on all genes of the network from Cluster A and from Cluster B, Supplementary Table 3 ^33, 34^. We found significantly enriched biological processes, molecular functions and cellular components based on Bonferroni corrected p-value for the case and control dominated networks. Prominent among the biological processes and molecular functions associated with preeclampsia were antigen processing and presentation, cellular movement (axon guidance and microtubules) and T cell receptor signaling.

We previously reported the Database for Preeclampsia (dbPEC) which archives a curation-based collection of genes associated with preeclampsia and their association with clinical features and concurrent conditions ^35^. We compared the genes from our univariate analysis and the genes from both case and control dominated layered networks to those in the database. We found two overlapping genes from the univariate gene list (TTN and CCL14) that were included in dbPEC. We also found three overlapping genes from the layered network of Cluster A (FN1, KIF2A, VCAM1). By over representation analysis, Cluster A is significantly enriched for genes previously shown to be associated with preeclampsia in dbPEC (p< 0.0033).

Aggregating information from an array of risk alleles and or genes, also known as a polygenic risk score (PRS), is a means to predict an individual’s phenotype or risk of disease based on their genomic profile.^36^ Gene-based models trained on the training set and tested on the test set with an LFDR threshold of 0.1 achieved the highest AUC for the ROC of 0.524. We hypothesized that our PPI network analysis would provide increased predictive power, and thus we also tested these models including the 13 unique genes identified in the preeclampsia dominant cluster. This resulted in an increase in the AUC to 0.57 with a 95% confidence interval between 0.383 and 0.732.

## Discussion

Preeclampsia is a life-threatening, multi-system hypertensive disorder of pregnancy, which complicates up to 15 % of US deliveries ^8, 36-38^. The incidence is increasing^38^. It is recognized as a leading cause of maternal and fetal morbidity and mortality worldwide ^36^. Preeclampsia is characterized by varying degrees of maternal symptoms including elevated blood pressure, proteinuria and fetal growth restriction ^39^. Many clinicians believe that preeclampsia, severe preeclampsia, and early vs late preeclampsia are different disorders ^40-42^. Previously, using bioinformatic methods, we showed that there are discrete gene sets associated with these different phenotypes of preeclampsia ^35^.

We performed whole exome sequencing on women with idiopathic early-onset preeclampsia with severe features and singleton births <34 weeks’ gestation and compared them to term controls with no family history of preeclampsia. We developed *Proteinarium*, a multi-sample, PPI analysis and visualization tool, to identify clusters of patients with shared protein-protein interaction networks ^43^. Using seed genes from each patient, *Proteinarium* built individual networks based on the STRING database. The similarities between individual PPI networks were evaluated using a distance metric for clustering the samples. We identified a single, significant cluster with a predominance of cases with early-onset, severe features of preeclampsia. We also identified a single control-dominated cluster. The separation test of the unique genes from case and control dominated clusters confirmed that the two subnetworks forming clusters A and B exist in the different regions of the interactome. These results support our hypothesis that the genetic architecture of complex diseases is characterized by clusters of patients that have variants in shared gene networks and provide insights into the genetics of severe preeclampsia.

Several of the unique genes from the case dominated network have very plausible mechanistic connections to preeclampsia. Laminin β2 (LAMB2) is a glomerular basement membrane (GBM) component, required for proper functioning of the glomerular filtration barrier. It has a role in proteinuria ^44^ and serum laminin levels in preeclamptic patients are significantly higher than those in normal pregnancy ^45^. Hypoxia-induced upregulation of Quiescin Sulfhydryl Oxidase 1 (QSOX1) and an elevation in intracellular H_2_O_2_ leads to increased apoptosis in the placentae of pregnancies complicated by preeclampsia ^46^. QSOX1 protein is found in circulating extracellular vesicles of both preeclampsia and healthy pregnant women ^47^. Fibronectin 1 (FN1) might promote the development of preeclampsia by modulating differentiation of human extravillous trophoblasts, as well as formation of focal adhesions ^48-50^. Vascular Cell Adhesion Molecule 1 (VCAM1) is involved in cellular adhesion and serum concentrations of sVCAM-1 are significantly elevated in both mild and severe preeclampsia ^51^. Invasion of maternal decidua and uterine spiral arteries by extravillous trophoblasts is required for establishment of normal placenta. Human trophoblast migration requires Rac Family Small GTPase 1 (RAC1) and Cell Division Cycle 42 (CDC42) ^52^. Lower levels were found in preeclampsia samples than in normal term pregnancy samples, and decline significantly in severe preeclampsia ^53^. Protein tyrosine kinase 2 (PTK2) (focal adhesion kinase) is differentially expressed in preeclampsia and reported as among the promising biomarkers for preeclampsia ^54^. In the case-dominated subnetwork we observed Kinesin Family Member 2A (KIF2A) which is upregulated in the preeclamptic placenta ^4^. Up-regulated genes in the preeclampsia placenta have been shown to be associated with the regulation of diverse cellular processes, including matrix degradation, trophoblast cell invasion, migration and proliferation ^4^.

There have been several sequencing efforts including whole genome, whole exome and targeted sequencing on an array of preeclampsia phenotypes from diverse populations ^37, 55-64^. There is no consensus among the published results in regards to associated genes and variants. Since preeclampsia is a complex, polygenic disease, the lack of a consensus among these univariate comparisons might be expected in these early-stage studies. Among the 20 genes identified in our univariate analysis, only Titin (TTN) was identified in prior studies ^55, 59^. Protein-altering mutations in TTN have been identified in patients with cardiomyopathy and women with preeclampsia are more likely to carry TTN mutations associated with idiopathic cardiomyopathy and peripartum cardiomyopathy ^59^. Additioinally, we found 2 genes, Major Histocompatibility Complex, Class II, DQ Alpha 1 (HLA-DQA1) and Inositol 1,4,5-Trisphosphate Receptor Type 1 (ITPR1) that were reported in previous studies of preeclampsia ^56, 62^. None of these overlapping genes were among the unique genes identified in the shared layered networks. Likewise, no overlapping variants or genes were found in a recent genome-wide association meta-analysis study investigating genetic predispositions associated with preeclampsia ^37^.

Our analysis allowed us to identify clusters of patients with shared PPI networks associated with severe preeclampsia. Within the significant clusters, there were unique imputed genes (RAC1, KIF5B, PTK2, KIF5A, FN1, QSOX1, ARF4, VCAM1, CDC42, KIF2A) that were not among the top 60 seed genes selected by genotype testing. Nonetheless, our approach allowed us to identify these influential genes in the mechanism(s) underlying preeclampsia that would not otherwise have been identified by whole genome univariate variant analysis.

We also examined the unique proteins of the network of the control dominated cluster. Proteins in this network are associated with the ubiquitination process. They may serve a role that confer resilience against preeclampsia ^65, 66^. Although there are studies showing a relationship with hypertension - ubiquitination process and pregnancy, this still needs further investigation ^66^.

Whole exome sequencing, combined with a novel, multi-sample network analysis, and careful phenotyping contributed to our discovery despite the relatively modest size of our study. Concepts developed from network theory suggest that related diseases involve proteins in similar neighborhoods of the interactome ^67^. Based on these concepts, we hypothesized that the genetic architecture of preeclampsia is described by subgroups of patients with variants in shared genes in specific networks and pathways. We identified a significant subgroup of cases with shared PPI networks associated with severe preeclampsia. We believe that the careful phenotyping resulted in the high percentage of subjects being successfully assigned to significant clusters and the ability to observe distinct separation between the case and control dominated clusters.

We used the Identified genes and their associated variants to generate a polygenic risk score. Of greatest importance, the unique genes in the case dominant cluster enhanced the predicted power of our polygenic risk score. These results compare favorably with results of others employing a similar approach ^68^. The recent meta-analysis of GWAS studies generated a polygenic risk score from different genomic elements with an odds ratio of 1.25 in prediction of preeclampsia ^37^.

### Strengths and Limitations

While we were not expecting each patient to appear in a significant cluster and our study included only a modest sample size, we identified a significant subgroup of patients with shared PPI networks associated with severe preeclampsia. In order to leverage the likelihood of genetics discovery, we focused exclusively on women with severe, early-onset preeclampsia. Our analysis was restricted to evaluation of genetic variants in the maternal genome only. Future studies including fetal and/or paternal data will enhance the likelihood of genetic discovery.

## Conclusion

Using our unique network analysis, we were able to identify subsets of patients with shared networks, thus confirming our hypothesis about the genetic architecture of preeclampsia. Strict phenotyping of both cases and controls improved the likelihood of identifying these otherwise difficult to find genetic associations. Our network analysis identified genes which were imputed from the interactome and these imputed genes provide insights for severe preeclampsia that may otherwise have not been identified. As such, these are important candidates to include in meta-analyses of genetic associations with preeclampsia. Inclusion of the unique genes identified in cluster associated with severe preeclampsia increased the predictive power of the polygenic risk score. These results provide promise to further our understanding the mechanism underlying complex diseases like preeclampsia.

## Supporting information

Computational methods_Suppl

## Data Availability

After acceptance of this manuscript by a peer-reviewed journal, data will be available to the public.

## Acknowledgements

We thank the Kilguss Research Core at Women & Infants Hospital and The Center for Computation and Visualization (CCV) at Brown University.

---

**Supplemental Table 1**. Unique genes from control dominated cluster (Cluster B). *Genes alphabetically ordered.

**Supplemental Table 2**. Significantly enriched biological processes, molecular functions and cellular components based on Bonferroni corrected p-value for case and control dominated networks.

