## Supplementary material for "Integrated Protein Network Analysis of Whole Exome Sequencing of Severe Preeclampsia": Computational methods_Suppl

### 1 Network Analysis

We hypothesized that the genetic architecture underlying complex disorders is best explained by subsets of patients with variants in shared networks and pathways sufficient to express the phenotype. To analyze our whole exome sequencing data, we developed *Proteinarium,* a multi-sample PPI analysis and visualization tool [1]. The top 60 genes, corresponding to the most significant, differentially abundant variants for each patient (ranked by genotype testing p value) were used as the seed genes for input into *Proteinarium*. *Proteinarium* was implemented with the minimum path length parameter set to 2, to include only those pathways in which seed proteins are connected directly to each other and/or via a single intermediary protein. We refer to these intermediary connecting proteins as imputed proteins. *Proteinarium* clusters the subjects based on similarities of their PPI networks using the Unweighted Pair Group Method with Arithmetic Mean (UPGMA) algorithm [2], outputting a dendrogram for visualization of the clusters. Statistical significance for each branch under the dendrogram is calculated by Fisher exact test comparing the abundance of cases and controls in each cluster relative to the total number of samples and their group assignment.

### 2 Network Separation Testing

Separation testing is a computational approach for determining the genetic similarity between diseases by comparing their protein-protein interaction networks from the interactome [3]. It compares the shortest distances between network proteins *within* each disease or network to the shortest distances *between* the disease networks. A positive separation score indicates that there is a physical separation between networks within the interactome. We computed the separation score between the networks of significant clusters identified by *Proteinarium* [1].

**3 Polygenic risk score calculation**

We generated phenotype specific polygenic risk scores, using methodology specific for whole exome sequencing data. We adapted the methodology outlined in Fabbri et al to calculate a score for all genes, weighting each variant by its frequency and functional score/pathogenicity[4]. Genotype calls were excluded if the quality score was <30 and variants were excluded if the call rate was less than 90% and if the heterozygote count was smaller than both homozygotes. We used EIGEN annotations as measures of a variant's pathogenicity. Rare variants were extracted from the sequence data based on a minor allele frequency (MAF) of 0.05. We used PLINK to clump common variants based on linkage disequilibrium and used the variant with the highest functional score to be used as the index variant [5]. We calculated a per sample per gene risk score separately for the rare and common variants, and combined these for an overall gene-based score. The overall distribution of these scores did not significantly differ between the cases and controls. 70% of the samples were used as a training set, and the remaining 30% were used as the test set. Using the training data, a correlation adjusted T (CAT) score was calculated for each gene and a local false discovery rate (LFDR), given a gene's CAT score was also calculated. We used various thresholds for LFDR to select predictors for the prediction model. Five-fold cross validation, where one-fifth of the training set left out in each round, was utilized to estimate parameters for the GBM prediction model. Performance of the model was estimated using the area under the curve (AUC) of the receiver operating characteristic (ROC).
