## Supplemental Table 1 for "Integrated Protein Network Analysis of Whole Exome Sequencing of Severe Preeclampsia"

**Supplemental Table 1.** Unique Genes from control dominated cluster (Cluster B). *Genes alphabetically ordered.

| Gene Name | Gene* | HGNC id | Cluster | Imputed |
|---|---|---|---|---|
| Aurora kinase A | AURKA | 11393 | B | Yes |
| Cbl proto-oncogene B | CBLB | 1542 | B | Yes |
| Cell division cycle 20 | CDC20 | 1723 | B | Yes |
| Cell division cycle 5 like | CDC5L | 1743 | B | Yes |
| Golgi reassembly stacking protein 1 | GORASP1 | 16769 | B | Yes |
| Kinesin family member 2C | KIF2C | 6393 | B | Yes |
| NEDD4 E3 ubiquitin protein ligase | NEDD4 | 7727 | B | No |
| P21 (RAC1) activated kinase 1 | PAK1 | 8590 | B | Yes |
| SMAD specific E3 ubiquitin protein ligase 2 | SMURF2 | 16809 | B | Yes |
| Tripartite motif containing 21 | TRIM21 | 11312 | B | Yes |
| Ubiquitin C | UBC | 12468 | B | Yes |
