## Supplemental Table 2 for "Integrated Protein Network Analysis of Whole Exome Sequencing of Severe Preeclampsia"

**Supplemental Table 2.** Significantly enriched biological processes, molecular functions and cellular components based on Bonferroni corrected p-value for case and control dominated networks. GO terms that are associated with case dominated networks were represented with “Cluster A” and with control dominated networks were represented with “Cluster B”.

| GO term ID | Definition | p value |
|---|---|---|
| | **Case dominated cluster (A)** | |
| **Biological Processes** | | |
| GO:0019886 | Antigen processing and presentation of exogenous peptide antigen via MHC class II | 2.09E-06 |
| GO:0007411 | Axon guidance | 3.75E-06 |
| GO:0007018 | Microtubule-based movement | 1.31E-03 |
| GO:0050852 | T cell receptor signaling pathway | 1.32E-03 |
| GO:0038096 | Fc-gamma receptor signaling pathway involved in phagocytosis | 1.19E-02 |
| GO:0031295 | T cell co-stimulation | 3.20E-02 |
| **Molecular Function** | | |
| GO:0003777 | Microtubule motor activity | 6.23E-06 |
| GO:0008017 | Microtubule binding | 2.36E-02 |
| GO:0019899 | Enzyme binding | 2.36E-02 |
| GO:0005088 | Ras guanyl-nucleotide exchange factor activity | 2.84E-02 |
| **Cellular Components** | | |
| GO:0005829 | Cytosol | 2.18E-05 |
| GO:0016020 | Membrane | 6.02E-04 |
| GO:0005871 | Kinesin complex | 1.33E-03 |
| GO:0008091 | Spectrin | 3.66E-02 |
| GO:0012507 | ER to Golgi transport vesicle membrane | 4.59E-02 |
| | **Control dominated cluster (B)** | |
| **Biological Processes** | | |
| GO:0050852 | T cell receptor signaling pathway | 1.29E-06 |
| GO:0019886 | Antigen processing and presentation of exogenous peptide antigen via MHC class II | 5.36E-05 |
| GO:0042787 | Protein ubiquitination involved in ubiquitin-dependent protein catabolic process | 1.09E-03 |

| | | |
|---|---|---|
| GO:0007062 | Sister chromatid cohesion | 3.08E-03 |
| GO:0007067 | Mitotic nuclear division | 1.73E-02 |
| GO:0031145 | Anaphase-promoting complex-dependent catabolic process | 2.54E-02 |
| **Molecular Function** | | |
| GO:0005515 | Protein binding | 6.13E-04 |
| **Cellular Components** | | |
| GO:0005829 | Cytosol | 1.59E-08 |
| GO:0005654 | Nucleoplasm | 2.84E-04 |
| GO:0005813 | Centrosome | 6.55E-03 |
| GO:0008091 | Spectrin | 3.12E-02 |
| GO:0012507 | ER to Golgi transport vesicle membrane | 3.73E-02 |
| GO:0005634 | Nucleus | 4.27E-02 |
| GO:0043234 | Protein complex | 4.75E-02 |
